# Factor H-related protein 3 (FHR-3) deposition in kidney allografts – localization and correlation with complement activation

**DOI:** 10.1101/2023.10.24.23297478

**Authors:** Felix Poppelaars, Nicole Schäfer, Anita H. Meter-Arkema, Shrey Purohit, Bernardo Faria, Mariana Gaya da Costa, V. Michael Holers, Mohamed R. Daha, Diana Pauly, Marc A. Seelen, Joshua M. Thurman

**Affiliations:** Department of Internal Medicine, Division of Nephrology, University Medical Center Groningen, University of Groningen, Groningen, The Netherlands; Department of Medicine, University of Colorado School of Medicine, Aurora, Colorado, USA; Department of Orthopedic Surgery, Experimental Orthopedics, Centre for Medical Biotechnology (ZMB), University of Regensburg, Regensburg, Germany; Nephrology R&D Group, Institute for Research and Innovation in Health (i3S), São João University Hospital Center, University of Porto, Porto, Portugal; Department of Anesthesiology, University Medical Center Groningen, University of Groningen, Groningen, the Netherlands; Department of Nephrology, University of Leiden, Leiden University Medical Center, Leiden, The Netherlands; Experimental Ophthalmology, University Marburg, Marburg, Germany

**Keywords:** Complement, kidney transplantation, Innate Immunity

## Abstract

**Introduction:** Factor H-related proteins (FHRs) have emerged as novel players in complement-mediated diseases, as they exhibit structural resemblances to factor H but lack the regulatory domains, enabling them to antagonize factor H and increase complement activation through several activities. Despite the widely importance of the complement system in kidney transplantation, FHRs have not been studied in this context. Utilizing a novel monoclonal antibody, we investigated the presence of FHR-3 in kidney allografts.

**Methods:** The RTEC-2 monoclonal antibody was validated using immunohistochemistry, Western Blot analysis, and immunoprecipitation combined with mass spectrometry. FHR-3 deposition, localization, and the relationships to complement activation were analyzed in human kidney biopsies obtained pre-transplantation from living and deceased donors, and post-transplantation in cases with acute tubular necrosis, acute cellular and vascular rejection, or chronic rejection.

**Results:** Glomerular FHR-3 deposition was detected in kidneys from deceased, but not living, donors before transplantation. Additionally, we observed FHR-3 deposition in post-transplant settings, both in cases of rejection and non-rejection. While tubular and vascular deposition of FHR-3 was observed in some cases, FHR-3 was predominantly seen in the glomeruli, where it was primarily localized to podocytes. Moreover, co-localization of FHR-3 and C3d was rarely detected, with most cases exhibiting separate and non-overlapping staining patterns for both antigens, However, there was a moderate correlation between the staining intensity of the FHR-3 and C3d in the kidney biopsies (*r*=0.38, P=0.01).

**Conclusion:** We detected FHR-3 deposition in kidney allografts under inflammatory conditions, primarily colocalizing with podocytes in both the presence and absence of complement activation.

## Introduction

The complement system is a defense system that in the context of kidney transplantation becomes hostile.^1,2^ Studies have shown that complement activation occurs in organ donors prior to transplantation, in kidney allografts during ischemia-reperfusion, and in the recipient during rejection.^2,3^ Renal allografts can be retrieved from living donors, and organ donors after brain death and circulatory death.^4^ In deceased organ donors, complement deposition is already observed in the kidney prior to transplantation.^2,3,5^ During transplantation, ischemia-reperfusion injury occurs due to an initial cessation of blood supply followed by a subsequent restoration of perfusion.^6^ Numerous studies have highlighted the vital role of the complement system in the pathophysiology of renal ischemia-reperfusion injury.^7,8^ Additionally, rejection is a major obstacle in renal transplantation and is divided into antibody-mediated rejection and cellular rejection.^9^ In antibody-mediated rejection, complement activation is triggered by donor-specific antibodies, while in cellular rejection, the complement system regulates the activity and differentiation of participating immune cells.^10,11^ Moreover, complement activation is linked to both short- and long-term adverse outcomes following kidney transplantation.^5,12,13^ Thus, complement activation is consistently observed in various settings before, during and after transplantation, all of which are indicative of pathological processes.

In health, complement activation is strictly controlled by various regulators such as Factor H.^14,15^ Factor H prevents unwanted complement activation by promoting C3-convertase dissociation, competing with factor B for C3b binding, and by acting as a co-factor for factor I-mediated conversion of C3b into iC3b.^16^ Its structure consists of 20 repetitive units known as short consensus repeat or complement control protein (CCP) domains, where the first four units are crucial for complement inhibition, and the last two units, along with a region in CCPs 6–8, are essential for ligand and cell surface recognition.^15,16^ Humans possess five genes adjacent to the complement factor H gene (*CFH*) that code for the factor H-related proteins (FHRs).^16,17^ The FHRs also consist of CCP domains and share structural similarities with the factor H domains involved in ligand and surface binding, but lack the regulatory domains.^18^ For example, domains 1 to 3 of FHR-3 share 91%, 85%, and 62% similarity with domains 6 to 8 in factor H, while domains 4 and 5 of FHR-3 exhibit 64% and 37% similarity to domains 19 and 20 of factor H.^18^ The current belief is that FHRs antagonize Factor H’s complement regulatory ability by competing for the binding to specific surfaces, leading to further complement activation due to their lack of regulatory capacity.^16,18,19^ Furthermore, more recent work has suggested that surface-bound FHRs can also directly promote complement activation by recruiting C3 to that surface.^20^ In accordance, mutations, genetic rearrangements, and duplications in FHRs genes (*CFHR*) have been associated with several renal diseases.^15,16^ Despite the critical role of complement in transplantation, there has been little inquiry into the role of FHRs in this context.

To study the role of FHRs as complement deregulators, the challenge lies in determining the *in vivo* interplay between FHRs and complement activation. Noteworthy, a major limitation has been the lack of specific monoclonal antibodies (mAb) that discriminate between the various FHRs and Factor H.^16,17^ Recently, a novel mAb against FHR-3 was described (RETC-2), showing promise for local and systemic detection without cross-reactivity with other FHRs or factor H.^21^ The average serum levels of FHR-3 are much lower than factor H, making it unlikely for any physiologically relevant competition to occur in the circulation between FHR-3 and the more abundant factor H, and instead suggesting a more plausible local role for FHR-3.^16,17,21,22^. The current study aims to investigate FHR-3 deposition in the kidneys during kidney transplantation settings characterized by complement activation, such as in deceased donors prior to transplantation and in transplanted kidneys with acute tubular necrosis and various forms of rejection. Additionally, we aimed to assess whether the presence of FHR-3 aligns with (enhanced) complement activation, examining whether FHR-3 acts as an amplifier of local complement activation in kidney transplantation.

## Materials and Methods

### Kidney Biopsies

Frozen kidney sections were obtained from two groups of deceased organ donors: brain dead organ donor and organ donors after circulatory arrest (n=5 per group) before organ retrieval. Additionally, we used frozen sections from human kidney biopsies with various conditions, including acute T cell– mediated rejection grades Banff IA, Banff IB, Banff IIA, Banff IIB, chronic rejection, and acute tubular necrosis (n=6 per group), in accordance with the defining criteria of the official Banff classification for rejection in kidney allografts.^23,24^ Furthermore, frozen sections from unaffected areas of kidneys with renal cell carcinoma, following surgical tumor excision, were included (n=6). As healthy controls, pre-implantation biopsies were collected from living kidney donors (n=8).

### Primary Antibodies

FHR-3 was detected using a mouse IgG2b kappa mAb to human FHR-3, clone RETC-2 (generated by the Regensburg Therapy Complement group, University Hospital Regensburg, Regensburg, Germany), which was previously shown to specifically detect FHR-3 without cross-reacting with factor H or any of the other FHRs.^21^ A mouse IgG2b kappa isotype was used as a control antibody (#14-4732-85, Invitrogen, Thermo Fisher Scientific). Complement deposition was detected using a rabbit polyclonal antibody (pAb) to human C4d (BI-RC4D, Biomedical), a rabbit pAb to human C3d (A0063, Dako), and a FITC-labeled mouse mAb to human C5b-9, clone aE11 (HM2167F, Hycult). Goat antiserum against human factor H (A312, Quidel) was used as a control for Western Blot analysis. Specific cell populations in the glomerulus were detected using a rabbit pAb to mouse CD31 that cross-reacts with human CD31 (ab28364; Abcam), a rabbit pAb to human platelet-derived growth factor receptor-β (PDGFR-β) (RB-1692, Thermo Fisher Scientific), a rabbit pAb to human podocin (P0372, Sigma-Aldrich), a goat pAb to human podocalyxin (AF1658, R&D systems), a rabbit pAb to human complement receptor 1 (CR1/CD35) (gifted by Mohamed R. Daha, generated previously in the laboratory of Nephrology, Leiden, The Netherlands), a goat pAb to human vimentin (AF2105, Novus Biologicals), and a pAb to human desmin (ab15200; Abcam).

### Immunofluorescence

The anti-FHR-3 mAb RTEC-2 stained frozen sections, but the antibody did not work for staining paraffin sections. Frozen human sections (4 μm thick) were air-dried, followed by fixation with cold acetone. Subsequently, they were blocked with 0.03% H_2_O_2_ in PBS. The anti-FHR-3 mAb RTEC-2 was incubated overnight at 4°C, and the binding was detected using a horseradish peroxidase (HRP)-conjugated goat anti-mouse IgG antibody (P0447, Dako), visualized with tetramethylrhodamine (TRITC) (SAT702001EA, TSA System). Nuclei were counterstained with 4,6-diamidino-2-phenylindole (DAPI). As a negative control, the anti-FHR-3 mAb was replaced with the isotype control or incubated without the secondary antibody. All antibodies were diluted in PBS with 1% bovine serum albumin (BSA). After each step, sections were washed three times for 5 minutes in PBS.

For double immunofluorescence staining, sections were incubated simultaneously with the anti-FHR-3 mAb and the antibody directed against either a complement protein or a cell surface marker. Antibody binding was detected using a mixture of secondary antibodies, including a goat anti-mouse IgG HRP-conjugated antibody and a goat anti-rabbit fluorescein isothiocyanate (FITC)-conjugated antibody (4050-02; Southern Biotech Technology), or a donkey-anti-goat FITC-conjugated antibody (6420-02; Southern Biotech Technology). HRP-conjugated secondary antibody binding was visualized using TRITC (SAT702001EA, TSA System). The exception was the double staining of FHR-3 with C5b-9 since the anti-C5b-9 mAb was already FITC-conjugated. In this case, we first incubated the anti-FHR-3 mAb, followed by the appropriate HRP-conjugated secondary antibody, and subsequently with the anti-C5b-9 mAb. Nuclei were counterstained with DAPI. To exclude cross-reactivity of the secondary antibodies with the other primary antibodies we performed the following controls: (i) the anti-FHR-3 mAb with both secondary antibodies, and (ii) the antibody directed against either a complement deposition or a cell surface marker with both secondary antibodies. None of the stainings showed cross-reactivity of the secondary antibodies with the other primary antibodies (data not shown).

### Cell Culture

Immortalized human podocytes were generously provided by Dr. Moin A Saleem.^25^ These cells exhibit undifferentiated proliferative growth at 33°C, while shifting to 37°C induces growth arrest and triggers differentiation into a parental podocyte phenotype. Undifferentiated cells were cultured at 33°C in RPMI 1640 medium supplemented with penicillin, streptomycin, insulin, transferrin, selenite, and 10% fetal calf serum.^25^ Upon reaching 70–80% confluence, they were transitioned to 37°C for differentiation, a process lasting 14 days. Podocytes were maintained in serum-free medium 24 hours prior to the addition of stimuli and throughout the experiment. In the experiments, we used recombinant human C3a (Hycult Biotech), recombinant human C5a (Hycult Biotech), and a combination of the two to stimulate the podocytes. After stimulation for 24 hours, both supernatants and podocytes were collected for western blot analysis.

### Western Blot

Recombinant human factor H and FHRs were produced in the Expi293 Expression System (Thermo-Fisher) and purified using a HisTrap HP column (GE Healthcare) as previously described.^26,27^ Native human factor H was purchased (A137, Complement Technology, Inc.). For human kidney samples (n=3), three 20 μm cryostat sections were lysed in RIPA buffer (1% NP40, 0.1% SDS, 10 mM β-mercaptoethanol) containing protease inhibitors. The samples were lysed on ice, followed by centrifugation at 16,000g for 15 minutes at 4°C, and the supernatants were collected. Protein concentration was determined using the Pierce Protein assay (Thermo Fisher Scientific) and 20 μg of total protein was used per kidney lysate sample. For immortalized human podocytes, cells were lysed in RIPA buffer and processed similarly to the human kidney lysates, except 10 μg of total protein was used for Western Blot analysis per cell lysate. Furthermore, in the analysis of the supernatant from immortalized human podocytes, 25 μl was utilized for Western blotting.

Samples were separated by SDS-PAGE on 4-20% gradient polyacrylamide gels using the BioRad electrophoresis system. The proteins were then electroblotted onto Nitrocellulose membranes, followed by immune detection. In brief, membranes were first blocked in 5% ELK/TBS with 0.05% Tween for 1 hour at room temperature, followed by an overnight incubation at 4°C with primary antibodies directed against FHR-3 or factor H in 3% BSA/TBS with 0.05% Tween. Subsequently, membranes were washed five times in TBS with 0.05% Tween and incubated with an appropriate secondary antibody conjugated to peroxidase in 5% BSA/PBS with 0.05% Tween. After five additional washes in TBS and 0.05% Tween, bound antibodies were detected using the ECL Western blotting detection system following the manufacturer’s protocols.

### Immunoprecipitation-Mass Spectrometry Antibody Validation

Five mg of tosylactivated dynabeads (Life Technologies) were either conjugated to 100 μg anti-FHR-3 mAb RETC-2, or 100 μg of the isotype control according to the manufacturer’s protocol. Next, 375 μg of total protein from a human kidney lysate of a deceased organ donor was incubated overnight with the mAb-coupled dynabeads on the roller bank at at 4°C. After removal of the supernatant, dynabeads were washed and the bound proteins were eluted in 50 µL using reducing Lemli sample buffer with dithiothreitol. Protein levels were determined using discovery-based proteomics (using label free quantification) for relative protein concentrations as previously described.^28^ Briefly, in-gel digestion was performed on an equivalent of 25 µL immunoprecipitation eluate using 150 ng trypsin (sequencing grade modified trypsin, Promega) after reduction with 10 mmol/L dithiothreitol and alkylation with 55 mmol/L iodoacetamide proteins.^29^

Discovery mass spectrometric analyses were performed on a quadrupole orbitrap mass spectrometer equipped with a nano-electrospray ion source (Orbitrap Exploris 480, Thermo Scientific). Chromatographic separation of the peptides was performed by liquid chromatography (LC) on a Evosep system (Evosep One, Evosep) using a nano-LC column (EV1137 Performance column 15 cm x 150 µm, 1.5 µm, Evosep; buffer A: 0.1% v/v formic acid, dissolved in milliQ-H2O, buffer B: 0.1% v/v formic acid, dissolved in acetonitrile). Half of the digests were injected in the LC-MS and the peptides were separated using the 30SPD workflow (Evosep). The mass spectrometer was operated in positive ion mode and data-independent acquisition mode (DIA) using isolation windows of 16 m/z with a precursor mass range of 400-1000, switching the FAIMS between CV-45V and -60V with three scheduled MS1 scans during each screening of the precursor mass range. LC-MS raw data were processed with Spectronaut (version 18.2.230802) (Biognosys) using the standard settings of the directDIA workflow except that quantification was performed on MS1. For the quantification, the Q-value filtering was setting to the classic setting without normalization or imputation.

### Scoring of the Kidney Biopsies

The biopsies were scored by two independent observers in a blinded fashion. For every section, a score ranging from 0 to 4+ was given for C3d and FHR-3, with a score of zero meaning there was no positive staining. Correlations were assessed by using Spearman’s correlation.

### Ethics

The use of human biopsies was approved by the Institutional Review Board (METc 2014/077) of the University Medical Center Groningen, Groningen, The Netherlands. Patient data were processed and stored according to the Declaration of Helsinki. Clinical and research activities followed the principles of the Declaration of Istanbul on Organ Trafficking and Transplant Tourism.

## Results

### Prominent Glomerular FHR-3 Deposition in Kidneys from Deceased Donors

Immunofluorescence analysis for FHR-3 was initially performed on kidney tissue obtained from different organ donors prior to transplantation, with liver tissue serving as the positive control for FHR-3. In the liver, hepatocytes exhibited positive staining for FHR-3 (Fig. 1A). Kidney biopsies from living donors obtained at the time of donation, with the kidney in situ and before clamping arterial and venous circulation, showed no FHR-3 staining (Fig. 1B), except for faint glomerular FHR-3 staining in 1 out of 9 cases. In contrast, positive staining was observed in the glomeruli of all kidneys from organ donors after circulatory arrest (Fig. 1C) and brain-dead organ donors (Fig. 1D). Additionally, FHR-3 deposition was occasionally seen on the apical side of some tubules to a lesser extent. Nuclear staining for FHR-3 was also observed in both liver and kidney tissue. No positive staining was detected when the anti-FHR-3 antibody was replaced with PBS or an appropriate isotype control (data not shown). The specificity of the mAb against FHR-3 was confirmed through Western Blot analysis using recombinant FHRs and factor H as well as native factor H, which produced a distinct signal for recombinant FHR-3 (Fig. 2A). In contrast, goat antiserum raised against human factor H was able to recognize all recombinant FHRs and factor H, as well as native factor H, to varying degrees (Fig. 2B). Furthermore, Western Blot analysis of renal lysates from three deceased donors with positive FHR-3 staining revealed distinct bands corresponding to the molecular weight of FHR-3 as a monomer in different glycoforms (between 35 – 55 kDa), along with an unknown band around 180 – 200 kDa (Fig. 2C). Neither bands were visible on Western Blot when the anti-FHR-3 antibody was replaced with PBS or an appropriate isotype control (data not shown). Lastly, we conducted immunoprecipitations using the anti-FHR-3 mAb, along with an isotype control, on a single kidney lysate, and analyzed the results using mass spectrometry. FHR-3 was exclusively identified in the immunoprecipitations performed with the anti-FHR-3 mAb, and not in those using the isotype control (Fig. 2D). In summary, these findings demonstrate the ability of the anti-FHR-3 mAb to consistently detect native FHR-3 in human kidneys through different techniques.

**Figure 1.**
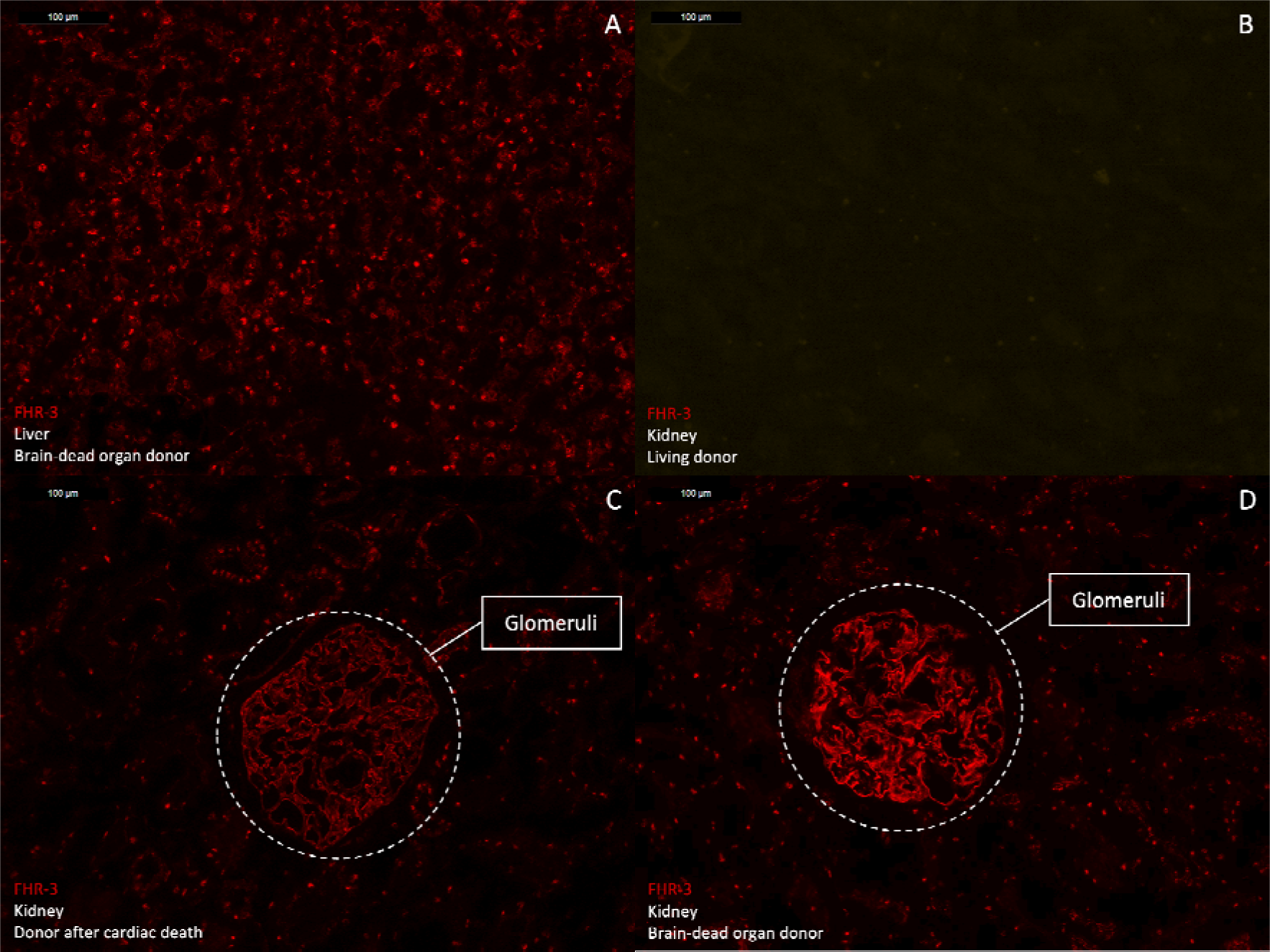
FHR-3 deposition in the glomeruli of kidneys from deceased donors. Representative images from Factor H-related protein 3 (FHR-3) staining using the monoclonal antibody RETC-2 on frozen tissue (n=3/per group). (A) Positive staining for FHR-3 in hepatocytes from a discarded liver of a deceased donor. (B) No staining for FHR-3 in kidney biopsies from living donors obtained at the time of donation, before retrieval while the kidney was in situ, and before clamping arterial and venous circulation. Glomerular staining for FHR-3 in kidney tissue from deceased organ donors (C) following circulatory arrest and (D) after brain death, before transplantation.

**Figure 2.**
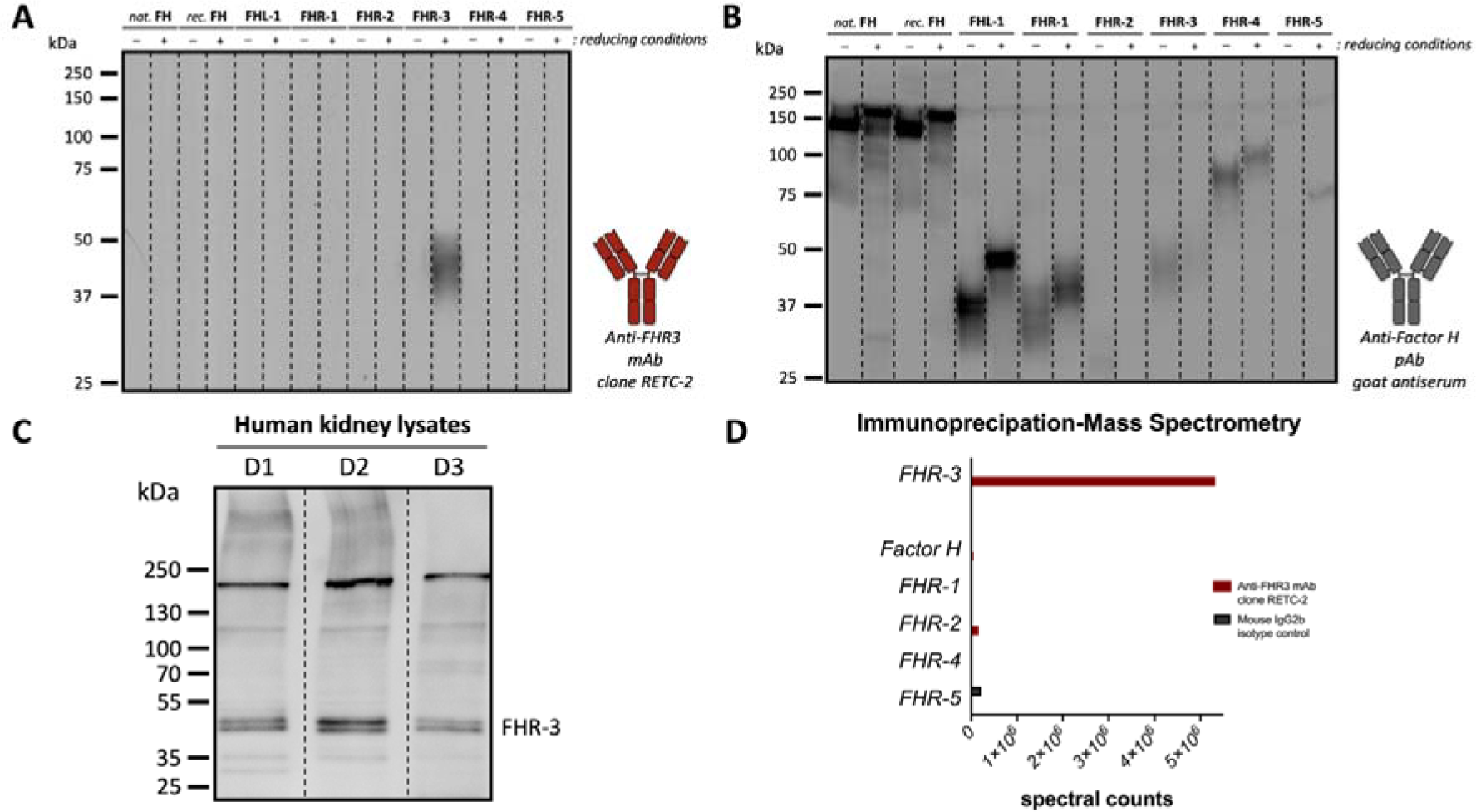
Specific detection of native FHR-3 in human kidneys with RETC-2 monoclonal antibody. Western blot analysis of recombinant factor H and factor H-related proteins (FHRs), as well as native factor H, confirmed that the (A) anti-factor H-related protein 3 (FHR-3) monoclonal antibody (mAb) RETC-2 exclusively recognizes recombinant FHR-3 but not other proteins of the factor H family. In contrast, (B) polyclonal goat antiserum against human factor H recognized all proteins to varying degrees. (C) The mAb RETC-2 detected clear protein bands corresponding to FHR-3 glycoforms ranging from 35 to 55 kDa in Western blot analysis of reduced human kidney lysates from three deceased donors, all of whom exhibited positive FHR-3 staining. (D) Immunoprecipitation on a single kidney lysate using the anti-FHR-3 mAb or the isotype control, combined with mass spectrometry, revealed that among the Factor H family, the RETC-2 mAb exclusively detects FHR-3.

*FHR-3 Deposition Partially Colocalizes with Complement Activation in Kidneys from Deceased Donors* Next, we assessed whether the presence of FHR-3 in kidneys from deceased donors co-localized with complement activation. Macroscopically unaffected areas of kidneys with renal cell carcinoma following surgical tumor excision were included as controls (Fig 3A – D). C3d was used as a marker for C3 activation since it is covalently bound to the surfaces. Double staining of FHR-3 and C3d revealed partial colocalization in the glomeruli, along with distinct non-overlapping distributions for each antigen (Fig. 3E – L). Some portions of the glomeruli were only positive for FHR-3, while tubules showed single positivity for C3d. Since FHRs have been suggested to interact with complement proteins in more proximal and terminal pathways ^30–33^, we extended our investigation to include double staining of FHR-3 with C4d and C5b-9 in kidneys from deceased donors. We observed partial colocalization in the glomeruli for C4d and FHR-3 (Fig. 4A – D), similar to what was seen for C3d and FHR-3 (Fig. 4E – H). In contrast, minimal to no co-localization was observed between FHR-3 and C5b-9, with C5b-9 being detected around tubules and deposited on the vasculature. (Fig. 4I – L). Collectively, these findings suggest that FHR-3 deposition in kidneys from deceased donors can occur both in the presence and absence of complement activation.

**Figure 3.**
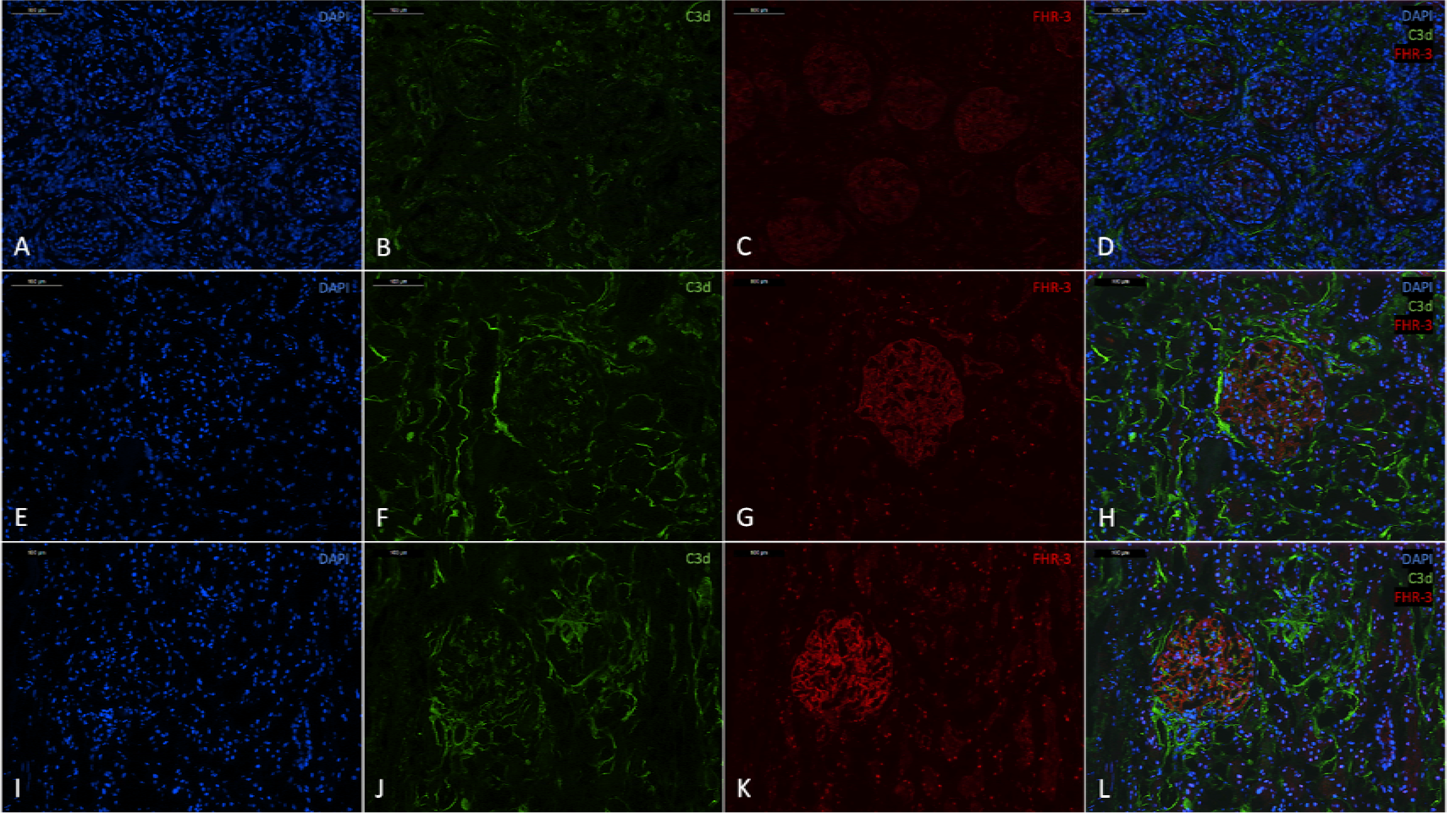
Partial glomerular colocalization of FHR-3 and C3d in kidneys from deceased donors. Representative images from immunofluorescent double staining of factor H-related protein 3 (FHR-3) and C3d in tissues obtained from (A-D) unaffected parts of a kidney following surgical tumor excision for renal cell carcinoma (RCC), (E-H) donor kidneys prior to transplantation from organ donors after circulatory arrest (DCD), and (I-L) donor kidneys prior to transplantation from organ donors after brain death (n=3/per group). Nuclei were counterstained with DAPI (blue in A, E, and I), C3d was stained with fluorescein isothiocyanate (FITC)-conjugated antibodies (green in B, F, and J), and FHR-3 was visualized using the tetramethylrhodamine (TRITC) (red in C, G, and K). Double stainings (D, H, and L) revealed partial glomerular colocalization of FHR-3 and C3d in kidneys from deceased organ donors prior to transplantation.

**Figure 4.**
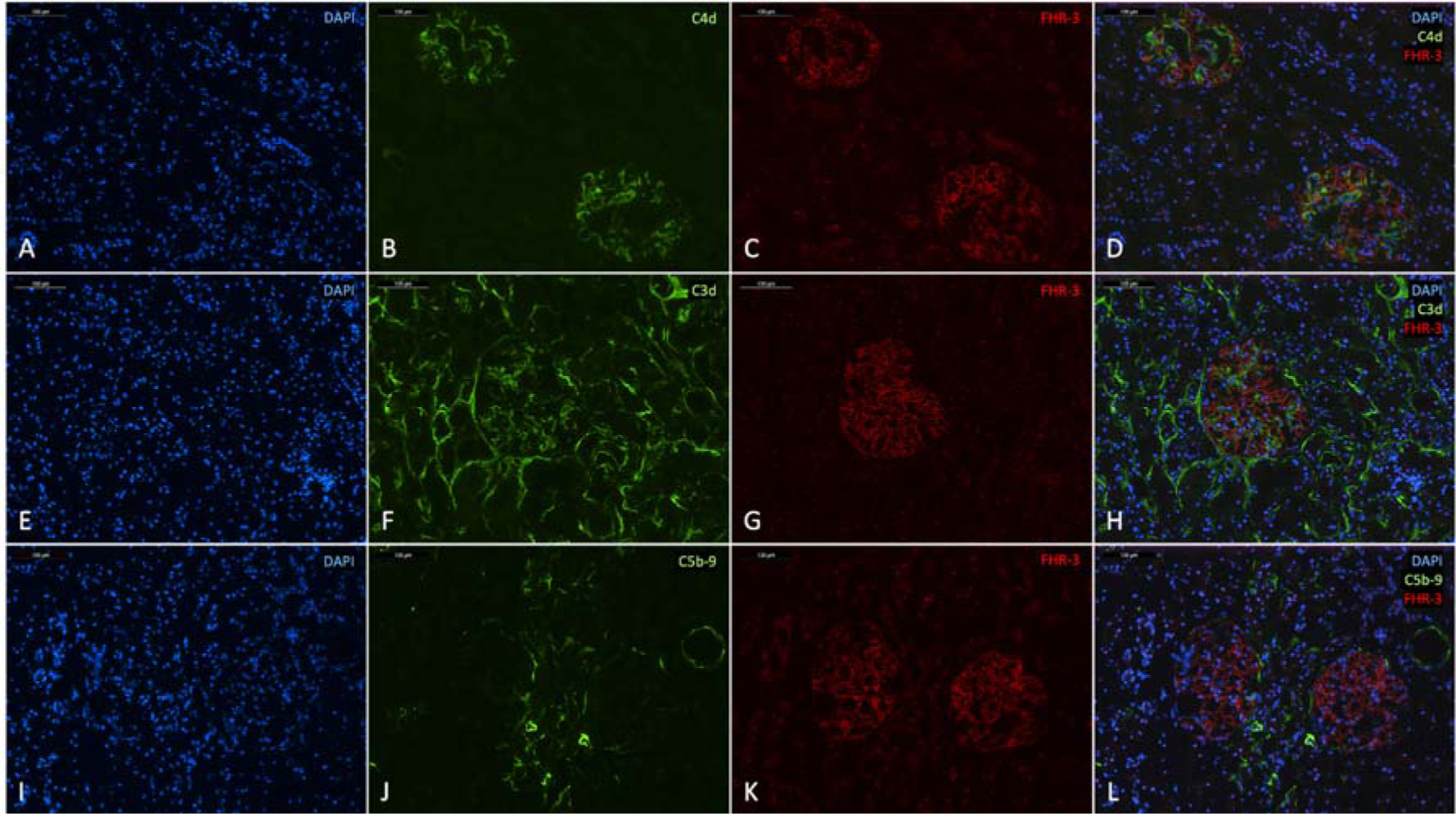
Staining patterns of FHR-3 and complement activation in kidneys from deceased donors. Representative images from immunofluorescent double staining of factor H-related protein 3 (FHR-3) with (A-D) C4d, (E-H) C3d, and (I-L) C5b-9 in kidney allografts prior to transplantation from organ donors after brain death (n=3 per staining). Nuclei were counterstained with DAPI (blue in A, E, and I). Complement deposition was stained with fluorescein isothiocyanate (FITC)-conjugated antibodies (green in B, F, and J), and FHR-3 was visualized using the TSA tetramethylrhodamine (TRITC) System (red in C, G, and K). Double stainings (D, H, and L) revealed partial glomerular colocalization of FHR-3 with C4d and C3d but not with C5b-9 in kidneys from deceased organ donors prior to transplantation.

*FHR-3 Deposition in the Kidneys Is Observed in Various Pathologic Post-Transplant Conditions* Subsequently, we analyzed the deposition of FHR-3 along with C3d in post-transplant biopsies diagnosed with acute tubular necrosis, acute cellular or vascular rejection (Banff IA, IB, IIA, IIB), and chronic rejection (Figure 5). In comparison to pre-implantation biopsies of living donors, renal transplant biopsies with acute tubular necrosis from patients clinically diagnosed with acute kidney injury exhibited strong intensity of glomerular FHR-3 deposition. Tubular FHR-3 deposition was also noted in 3 out of 6 cases (Fig. 6A). Notably, evident glomerular FHR-3 deposition was present in all cases of tubulointerstitial rejection (Banff IA, IB), while no or only faint glomerular FHR-3 staining was observed in cases of vascular rejection (Banff IIA, IIB). In contrast, vascular FHR-3 deposition was observed in 4 out of 6 cases with severe tubulitis (Banff IB, Fig. 6B), but was rare in cases with moderate tubulitis (1 out of 6 Banff IA cases) or vascular rejection (1 out of 12 Banff IIA and IIB cases). In kidney transplant biopsies with chronic rejection, positive FHR-3 staining was observed in the glomeruli, tubules, and vasculature in all cases (Fig. 6C).

**Figure 5.**
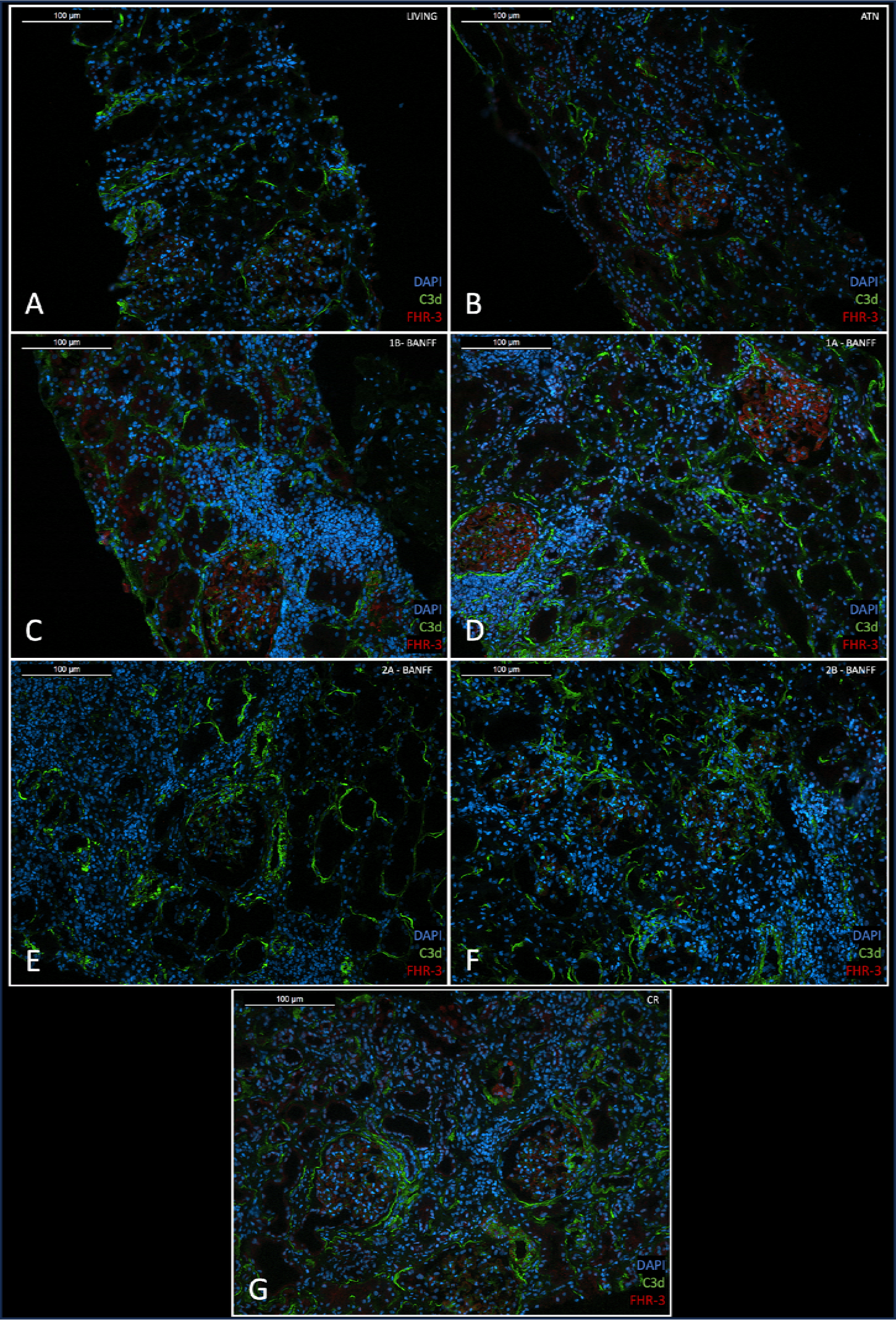
FHR-3 and C3d deposition in biopsies from living kidney donors and transplanted kidneys. Representative images from immunofluorescent double staining of factor H-related protein 3 (FHR-3 in red) and complement activation (C3d in green) in tissues obtained from (A) living donor kidneys (n=8), (B) acute tubular necrosis (ATN) (n=6), (C) Banff IA (n=6), (D) Banff IB (n=6), (E) Banff IIA (n=6), (F) Banff IIB (n=6), and (G) chronic rejection (CR) (n=6). Nuclei were counterstained with DAPI.

**Figure 6.**
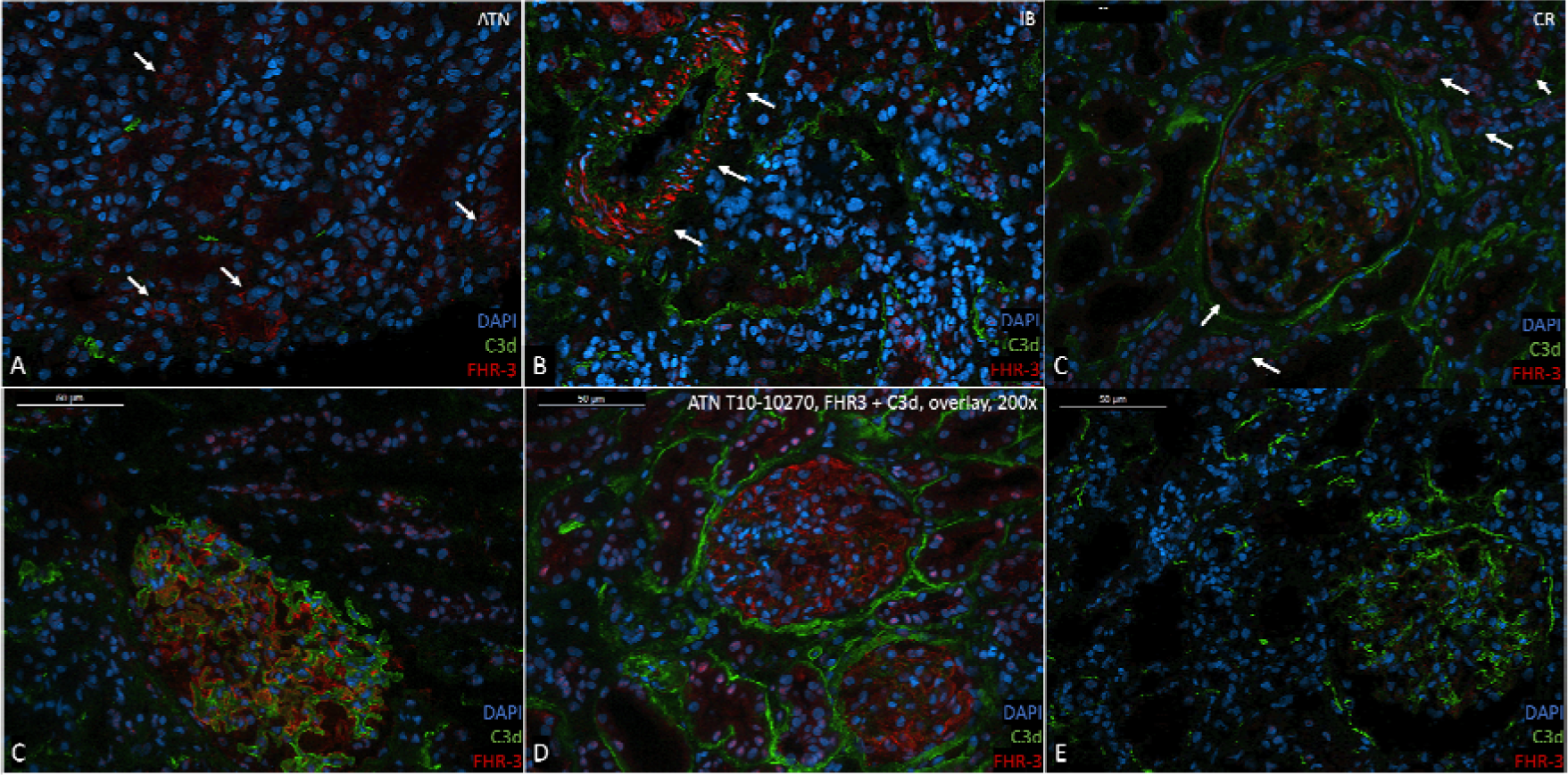
Diverse staining patterns of FHR-3 and C3d in biopsies from transplanted kidneys. Different staining patterns of factor H-related protein 3 (FHR-3 in red) and complement activation (C3d in green) in transplanted kidneys. Nuclei were counterstained with DAPI. Representative images of (A) tubular FHR-3 staining (arrows) in acute tubular necrosis (ATN), (B) vascular FHR-3 staining (arrows) in T-cell-mediated rejection with severe tubulitis (Banff IB), and (C) low-intensity FHR-3 staining in Bowman’s capsule (arrows) and tubules (arrows) in chronic rejection (CR). Exemplary images of double staining for FHR-3 (red) and C3d (green) in transplanted kidneys revealing (C) predominant positive glomerular staining for FHR-3, (D) glomerular co-localization of FHR-3 and C3d, and (E) predominant positive glomerular staining for C3d.

C3d deposition was also observed in kidney biopsies in various post-transplant conditions. Although renal C3d deposits were seen in pre-implantation biopsies of living donors, the intensity was notably lower, primarily localized in Bowman’s capsule and, to some extent, around the tubules. Furthermore, clear glomerular colocalization of FHR-3 and C3d was rarely observed (Fig. 6D). Instead, in most cases, positive staining for both antigens tended to exhibit a distinct, non-overlapping distribution. Across the biopsies, some displayed single-positive glomeruli staining for FHR-3 (Fig. 6E), while others exhibited single-positive staining for C3d (Fig. 6F). To examine the relationship between FHR-3 and C3d deposition, we quantified the staining intensity of both antigens in living donors and post-transplant conditions (Fig. 7A, Supplementary data Table S1). Despite the lack of co-location in most biopsies, C3d staining intensity did have a moderate correlation with FHR-3 staining intensity in the biopsies of kidney allografts overall (Fig. 7B, r = 0.38, P-value = 0.01). Once again, these findings indicate FHR-3 deposition in kidney allografts during inflammatory conditions occurs in the presence and absence of complement activation.

**Figure 7.**
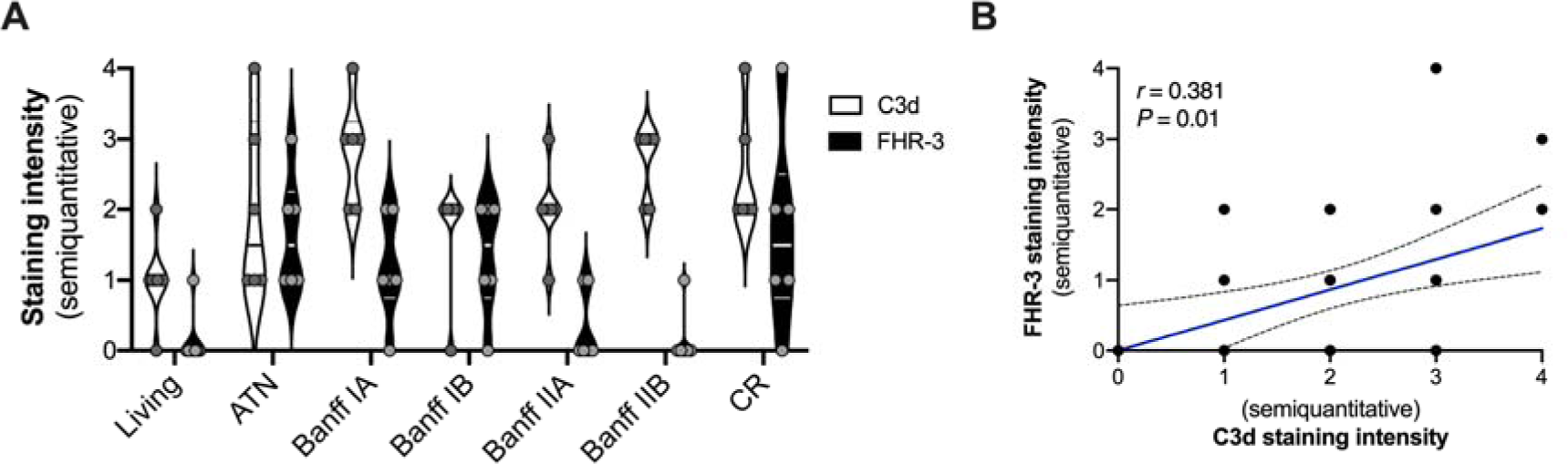
Quantification and correlation of FHR-3 and C3d staining in kidney allografts. (A) Quantification of staining intensity of factor H-related protein 3 (FHR-3) and complement activation (C3d) by double immunofluorescent in living donor kidneys, acute tubular necrosis (ATN), Banff IA, Banff IB, Banff IIA, Banff IIB, and chronic rejection (CR). Each biopsy was scored by two independent observers in a blinded fashion. (B) The correlation between the staining intensity of FHR-3 and C3d using the Spearman Rank correlation coefficient (r represents the spearman’s rho). The dashed blue lines show the 95% confidence interval for the regression line (blue).

### FHR-3 Staining Is Confined to Podocytes in the Glomeruli

Finally, we determined the localization of FHR-3 within the glomeruli since this was the primary site of deposition. The FHR-3 staining pattern under morphological examination hinted at podocytes, identifiable by their abundant cytoplasm, protrusions, and large nuclei. To substantiate this, we conducted double staining with various podocyte markers, including podocalyxin, podocin, CR1, vimentin, and desmin. Immunofluorescence staining of glomeruli from deceased donors affirmatively demonstrated FHR-3 presence on podocytes. FHR-3 double staining with podocalyxin (Fig. 8A – C), and podocin (Fig. 8D – F) showed significant overlapping reactivity. Podocin showed a more complete overlapping signal, suggesting that FHR-3 was localized in the slit diaphragms, rather than at the apical surface of the podocytes. While FHR-3 seems to colocalize in the same cells as vimentin (Fig. 8G – I) and desmin (Fig. 8J – L), components of the podocyte cytoskeleton, a substantial overlap in signaling was notably absent. This could suggest that while these intermediate filament proteins reside intracellularly, FHR-3 might be predominantly positioned on the cell membrane. Surprisingly, the CR1 staining did not overlay with FHR-3 (Fig. 8M – O). Finally, no signal overlap was seen for FHR-3 with the mesangial marker PDGFR-β or the endothelial marker CD31 (data not shown). To explore the ability of podocytes to produce FHR-3, we examined protein expression by immortalized cultured human podocytes. However, western blot analysis of both supernatants and cell lysates from human podocytes under baseline conditions and subsequent stimulation with anaphylatoxins did not reveal any production of FHR-3 at the protein level (data not shown). In conclusion, FHR-3 deposition is observed on podocytes, but the origin and mechanism of FHR-3 remains uncertain.

**Figure 8.**
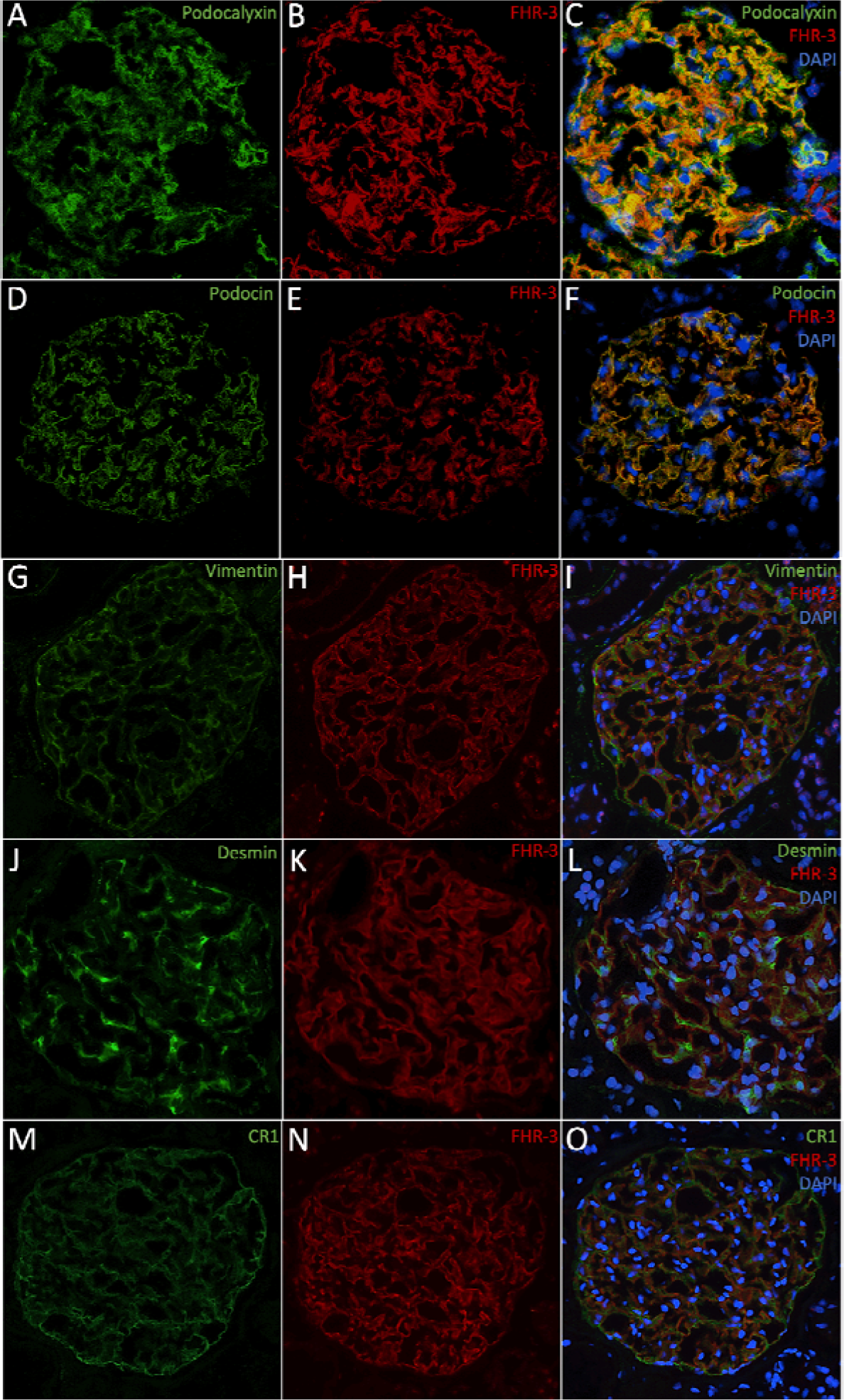
Glomerular FHR-3 localizes to podocytes in kidney allografts. Representative images from immunofluorescent double staining of factor H-related protein 3 (FHR-3) with (A-C) Podocalyxin, (D-F) Podocin, (G-I) Vimentin, (J-L) Desmin, and (M-O) complement receptor 1 (CR1) in kidney allografts prior to transplantation from organ donors after brain death. Nuclei were counterstained with DAPI (blue). Podocyte markers (green) were stained with fluorescein isothiocyanate (FITC)-conjugated antibodies (A, D, G, J and, M), and FHR-3 (red) was visualized using the TSA Tetramethylrhodamine (TRITC) System (B, E, I, K, N).

## Discussion

The importance of the complement system in the pathophysiology of kidney disease and kidney transplant failure is well-established.^1,2,15^ Recently, the FHRs have emerged as novel players in diseases characterized by complement dysregulation, with growing evidence indicating their antagonistic relationship with factor H and other means of promoting local activation.^16,17,20^ Since certain FHRs, like FHR-3, have significantly lower systemic concentrations compared to factor H, it has been hypothesized that FHRs may have a local role in complement-mediated diseases, rather than a systemic one.^21,22,34^ To investigate this further, it is imperative to gain a detailed understanding of the FHRs in tissue during disease, and to study their interaction with and relationships to complement activation. However, a significant obstacle has been the lack of specific antibodies due to the high structural homology between the FHRs and factor H.^16,17^ In this study, we used a recently developed specific mAb RTEC-2 against FHR-3.^21^ Our findings reveal that FHR-3 deposition occurs in kidney allografts under inflammatory conditions but is absent under native conditions. Furthermore, we determined that FHR-3 deposition is primarily localized on podocytes. Notably, we observed FHR-3 in kidney allografts, both in the presence and absence of colocalization with complement activation. This raises intriguing questions about the *in vivo* ability of the FHR-3 to bind to cell surfaces through both complement activation fragments and other ligands.

To our knowledge, this study represents the first comprehensive investigation into the immunohistological localization of FHR-3 in human renal biopsy specimens. Research on FHRs in kidney biopsies has been relatively limited. However, the available data suggest that deposition of FHRs may serve as a potential clinical biomarker, reflecting underlying tissue changes and holding promise for assessing the severity and predicting the progression of kidney disease.^35–38^ We observed glomerular FHR-3 deposition in deceased donor kidneys before transplantation, while FHR-3 was absent in living donor kidneys, which are typically considered ideal healthy controls in terms of quality. These findings align with proteomic analyses of deceased donor kidneys, confirming the presence of FHR-3.^39^ Moreover, Kaisar *et al.* recently demonstrated that in deceased donor kidneys activation, there is significant activation of proteolytic processes, leading to specific alterations in the podocyte cytoskeleton, which is associated with adverse posttransplant outcomes.^40^ We propose that FHR-3 deposition on podocytes arises from pathophysiological alterations within the deceased organ donor (e.g. prolonged ischemia and systemic inflammation) that damages the integrity of the glomerular filtration barrier and podocytes.^2,41,42^ Interestingly, In deceased donor kidneys undergoing normothermic machine perfusion, proteomic analysis unveiled an initially high abundance of FHR-3 in both the kidneys and their urine, which gradually diminished over time as the kidney’s metabolic activity was restored.^39^ These data further support the notion that FHR-3 presence is closely associated with inflammatory responses and tissue injury. Accordingly, we also identified the presence of FHR-3 in post-transplantation in kidney biopsies with acute kidney injury or different forms of rejection. FHR-3 staining was most prominent in cases of acute tubular necrosis and tubulointerstitial rejection, suggesting that tubular injury may be a contributing factor to the deposition of FHR-3. A recent study examining genome-level mismatches in kidney transplant donor-recipient pairs unveiled that a donor-recipient mismatch in the common gene deletion involving FHR-3 and FHR-1 (Δ*CFHR3,1*) was associated with the risk of acute rejection.^43^ However, these findings necessitate validation in larger cohorts and further substantiation through functional studies of FHR-1 and FHR-3 in the context of rejection.

In our study, we were unable to determine if FHR-3 originated from the kidney or the circulation. While we did not observe FHR-3 production in a podocyte cell line, it’s important to note that this does not rule out the possibility of local production by the kidney. Although the liver is conventionally regarded as the primary source of complement proteins, the kidney has been demonstrated to produce various complement factors thereby contributing substitutionally.^44,45^ Notably, FHR-3 has been detected in the perfusate of isolated deceased donor kidneys,^46^ while serum levels of FHR-3 were found to be elevated in deceased donors compared to healthy controls, suggesting that the two sources may not be mutually exclusive.^47^ Further studies are therefore needed to uncover the mechanisms that result in renal FHR-3 deposition. Like other FHRs, FHR-3 has been shown to bind different C3 activation fragments, competing with factor H.^21,48,49^ Although FHR-3 deposition was predominantly observed in kidney allografts with increased complement activation in our study, co-localization with C3 activation fragments was rare. Conversely, FHR-3 has also demonstrated to bind other ligands, such as glycosaminoglycans, as well as to malondialdehyde-epitopes on cellular debris and necrotic cells.^21,48,50–52^ In summary, our results suggest that the deposition of FHR-3 in kidney allografts is likely influenced by other ligands in addition to complement activation fragments.

While our understanding of the FHRs is still in its early stages, their significance in various diseases has been repeatedly demonstrated through unbiased genetic studies.^16,17^ In the case of IgA nephropathy, the presence of the ΔCFHR3,1 has been associated with a protective effect.^53–58^ This deletion is linked to reduced glomerular C3 deposition, decreased systemic C3 activation, and elevated systemic factor H levels in IgA nephropathy patients.^57–59^ Intriguingly, the Δ*CFHR3,1* is associated with a decreased risk of developing IgA nephropathy and age-related macular degeneration but carries an increased susceptibility to systemic lupus erythematosus and atypical hemolytic syndrome.^53,60–62^ These opposing associations highlight the context-dependent roles of FHRs, suggesting that these proteins are protective in specific scenarios (such as facilitating the removal of cellular debris and preventing autoantibody formation) while being harmful in other contexts (such as during complement activation on extracellular matrices and particular cell types). Recent findings have suggested that FHR-3 may have non-canonical roles, such as impacting cell viability.^63^ Whether FHR-3 plays a protective role in kidney transplantation by facilitating the removal of cellular debris and reducing alloimmunity or contributes harmfully by enhancing complement activation and inflammation remains to be determined.

The present study has a few limitations that should be taken into consideration. Firstly, it is an observational study with a relatively small sample size, which limits our ability to conduct a comprehensive analysis of the impact of FHR-3 deposition on clinical outcomes. Secondly, we lack genetic data regarding the copy variant numbers of FHR-3 and the Δ*CFHR3,1*, which are known to significantly influence circulating levels.^22,34^ Nevertheless, the absence of FHR-3 deposition in living donors, along with evident FHR-3 staining patterns in deceased donor kidneys and certain post-transplantation conditions, suggests that the observed results are unlikely to be solely attributed to differences in FHR-3 genotypes. Thirdly, we only explored FHR-3 production under limited conditions in podocytes and did not examine other renal cell types. However, this study has several key strengths, including (i) the inclusion of multiple groups covering both pre- and post-transplant conditions along with appropriate controls, (ii) the extensive validation of the specific FHR-3 monoclonal antibody, and (iii) the comprehensive analysis of FHR-3 with complement activation and different glomerular cell types.

In conclusion, this study has revealed the deposition of FHR-3 in kidney biopsies before and after transplantation, particularly under inflammatory conditions, with a predominant presence on podocytes. Furthermore, our observations showed only sporadic co-localization of FHR-3 with complement activation. These findings suggest the presence of FHR-3 on injured glomeruli in vivo, possibly irrespective of C3 activation fragments.

## Data availability statement

Data can be made available upon a reasonable request to the corresponding author.

## Abbreviations

ΔCFHR3,1: Gene deletion of FHR-3 and FHR-1
ATN: Acute tubular necrosis
BSA: Bovine serum albumin
CCP: Complement control protein domains
*CFH*: Complement Factor H gene
*CFHR*: Complement Factor H-related protein gene
CR: Chronic rejection
CR1: Complement receptor 1
DAPI: 4,6-diamidino-2-phenylindole
DBD: Donation after brain death
DCD: Donation after circulatory arrest
FHR-1: Factor H-related protein 1
FHR-3: Factor H-related protein 3
FHRs: Factor H-related proteins
HRP: Horseradish peroxidase
PDGFR-β: Platelet-derived growth factor receptor-β
pAb: Polyclonal antibody
mAb: Monoclonal antibody
TRITC: Tetramethylrhodamine

## Acknowledgements

We thank Marcory C.R.F. van Dijk, Maaike B. van Werkhoven, and Jeffrey Damman for their efforts and support in tissue collection. We also thank Jan-Luuk Hillebrands, Jaap van den Born, and Wendy Dam for their helpful discussions and assistance, as well as for sharing reagents for the staining of glomerular cell markers.

## Author Contributions

FP, DP, MAS, and JMT developed the concept, designed the study, and/or planned the experiments. NS and DP developed the FHR-3 antibody. FP, AHM, and MG conducted the experiments. FP, NS, AHM, DP, MAS, and JMT analyzed and discussed the data. NS, SP, BF, MRD, VMH, DP, MAS, and JMT provided materials. FP wrote the manuscript. All authors participated in manuscript discussions, offered feedback, contributed to the editing process, and approved the final version.

